# HALP and mHALP as Effective Tools for 90-Day Mortality Prediction in Heart Failure

**DOI:** 10.1101/2025.05.22.25328139

**Authors:** Abdulla Hourani, Arman David Sürmeli, Sai Keertana Devarapalli, Michał Oręziak

## Abstract

**Background:** Congestive heart failure (CHF) is a chronic syndrome with high mortality rates, necessitating risk stratification tools. Existing models are complex and may not incorporate crucial inflammatory and nutritional biomarkers. The Hemoglobin, Albumin, Lymphocyte, and Platelet (HALP) score and modified HALP (mHALP) score, which reflect nutritional status and systemic inflammation, emerged as potential prognostic tools. This study aimed to evaluate the predictive performance of HALP and mHALP scores for 90-day mortality in CHF patients.

**Methods:** This retrospective study utilized the MIMIC-IV database for 1000 adult CHF patients. HALP was calculated as [Hemoglobin (g/L) * Albumin (g/L) * Lymphocytes (/L)] / Platelets (/L), and mHALP by multiplying these components. The primary outcome was 90-day all-cause mortality. Univariate and multivariate logistic regression analyses were performed, and models were assessed using Receiver Operating Characteristic (ROC) analysis and Youden’s Index.

**Results:** Non-survivors (13.2%) had significantly lower median HALP (19.69 vs. 27.18) and mHALP (583,920.75 vs. 1,151,898.94) scores compared to survivors. In univariate analysis, HALP had an AUC of 0.63 and an OR of 0.25 for 90-day mortality, while mHALP showed a higher AUC of 0.68 and an OR of 0.22. After confounder adjusting, both log-transformed HALP (OR: 0.44) and mHALP (OR: 0.57) remained significant predictors of mortality. mHALP demonstrated better overall predictive performance.

**Conclusion:** Both HALP and mHALP scores are significantly associated with 90-day mortality in CHF patients, with mHALP exhibiting superior prediction. Easily calculable scores, derived from routine laboratory tests, offer a cost-effective, early risk stratification in CHF, potentially aiding in tailoring interventions.

## Introduction

Congestive heart failure (CHF) is a chronic and complex syndrome characterized by the heart’s inability to pump blood as efficiently as it should. According to data from the National Health and Nutrition Examination Survey (NHANES) 2017-2020, around 6.7 million (2.3%) of Americans greater than 20 years old had CHF, with the prevalence projected to increase by 46%, affecting >8 million people in 2030 [1]. Some well-established risk factors of CHF include ischemic heart disease, hypertension, diabetes, dyslipidemia, obesity, older age, and family history. Despite advances in treatment, prognosis remains poor, and mortality remains high. In 2020, CHF was a contributing cause of death in at least 415,922 Americans, accounting for around 1 out of 8 registered deaths [2]. Hence, early identification and timely intervention of high-risk patients are crucial for improving global health outcomes related to CHF.

Over the past few decades, researchers have developed various risk scores and predictive models to simplify diagnosis and enhance treatment strategies. The Seattle Heart Failure Model, MAGGIC Risk Score, and the CHARM score are among the widely used tools for predicting survival outcomes in patients with CHF [3-5]. While these models are robust in predicting mortality, they depend heavily on demographic factors and comorbidities—data that is not always reliably obtained due to incomplete clinical histories and inconsistencies in self-reported or documented information. Moreover, gathering necessary variables can be cumbersome and expensive, and the computation of these scores typically involves complex calculations. Additionally, these scores fail to incorporate important inflammatory and nutritional biomarkers that demonstrate significant prognostic value in CHF. There is a need for mortality risk scores that are easily accessible, reliable, inexpensive, and simple to use, in acute and chronic settings.

Routinely tested laboratory parameters such as hemoglobin, albumin, lymphocyte count, and platelet count have proven to be significant predictive factors of CHF mortality [6-9]. In the last decade, the Hemoglobin, Albumin, Lymphocyte, and Platelet score (HALP) has emerged as a novel prognostic tool for several types of cancers [10]. Hemoglobin and albumin indicate the nutritional status of a patient, while lymphocytes and platelet count give information about systemic inflammation. Since nutrition and inflammation play a key role in the disease progression, complications, and mortality associated with CHF, the HALP score can provide useful insights about a patient’s prognosis. Recent studies have analyzed the effectiveness of the HALP score as a tool to predict mortality in various clinical settings other than cancer, including ischemic stroke [11], STEMI [12], and proximal femur fractures [13]. However, research on the use of HALP score to assess mortality in CHF patients is currently limited.

Another study hypothesized that low platelets are a significant risk factor for mortality in patients with acute heart failure and developed the modified HALP (mHALP) score [14]. In this formula, platelets are multiplied with the other components of HALP. The authors found mHALP to be a significant predictor of mortality in these patients. The aim of this study is to evaluate the predicting performance of HALP and mHALP scores for 90 day mortality in CHF patients.

## Methods

### Data Source

This study utilized data from the Medical Information Mart for Intensive Care IV (MIMIC-IV) database, an extensive and publicly available electronic health record (EHR) repository maintained by the Beth Israel Deaconess Medical Center (BIDMC) in Boston, Massachusetts [15]. MIMIC-IV contains detailed clinical data, including demographics, laboratory results, medications, procedures, and mortality outcomes for patients admitted to the hospital and intensive care units (ICUs) between 2008 and 2022. Additionally, this study leveraged data from the MIMIC-Note module, which provides detailed clinical narratives, including discharge summaries and radiology reports. The combined utilization of MIMIC-IV and MIMIC-Note enabled robust phenotyping of patients with CHF and facilitated a comprehensive analysis of clinical, laboratory, and outcome data. This study was conducted following the Declaration of Helsinki and approval of the Institutional Ethics Board was not required due to the nature of the data, being anonymised and retrospective. The MIMIC-IV database is de-identified, ensuring patient confidentiality and privacy.

Patients were identified using ICD-9 and ICD-10 codes specific to CHF, with additional support from discharge summaries to improve case identification accuracy and extract ejection fraction and ST changes. Clinical notes were analyzed using ClinicalBERT [16], a transformer-based language model pre-trained on clinical notes from MIMIC-III, supported by regex-based methods to extract and verify CHF diagnoses and ejection fraction categories from discharge notes. To construct the study cohort, strict inclusion and exclusion criteria were applied. Adult patients with a primary diagnosis of CHF were included if they had complete clinical and laboratory data necessary for the analysis, given that laboratory tests are taken within 24h of admission. Patients with moderate or severe liver disease, immunosuppressant treatment, hematological cancer, immunodeficiency, or end-stage renal disease (eGFR < 15 mL/min/1.73m^2^) were excluded to minimize confounding and enhance the specificity of CHF-related outcomes. Additionally, patients with implausible laboratory values, including platelet counts <20 or >900 10^9/L, albumin <10 or >60 g/L, lymphocyte counts <0.2 or >10 10^9/L, and hemoglobin <50 or >250 g/L, were excluded to ensure data validity and reliability. To ensure the independence of observations, only the first hospital admission was retained. Patients aged over 89 years were excluded as their age was shifted in the original dataset to 300 to comply with HIPAA regulations.

### Variables

The primary predictors of interest were the Hemoglobin-Albumin-Lymphocyte-Platelet (HALP) score and its modified version (mHALP). It was calculated using the formula:

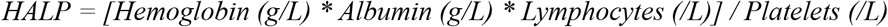

The modified HALP (mHALP) score was also calculated using the alternative formula:

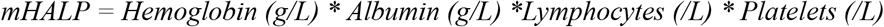

To adjust for potential confounders, several demographic, clinical, and laboratory covariates were included in the analysis. Demographic variables comprised age, gender, race, and marital status. Clinical variables included comorbidities such as hypertension, diabetes, chronic kidney disease, chronic obstructive pulmonary disease, history of myocardial infarction, and atrial fibrillation. Comprehensive laboratory data were also extracted, including white blood cell (WBC) count, total protein, creatinine, blood urea nitrogen (BUN), glucose, NT-proBNP, as well as liver function tests (AST, ALT, total bilirubin, and alkaline phosphatase).

### Outcome

The primary outcome of interest was all-cause mortality within 90 days of hospital admission, measured as a binary variable indicating survival or death.

### Statistical Analysis

Descriptive statistics were employed to summarize baseline characteristics, stratified by 90-day mortality. Continuous variables were presented as mean and standard deviation for normally distributed data or as median and interquartile range for skewed distributions. Categorical variables were summarized using frequencies and percentages. To compare differences between survivors and non-survivors, the Student’s t-test or Mann-Whitney U test was used for continuous variables, depending on the normality of distribution, while the Chi-squared test or Fisher’s exact test was applied for categoricals. Normality was rigorously assessed using the Anderson-Darling test for skewness and kurtosis, the Kolmogorov-Smirnov test for distribution comparisons, and visual inspection of histograms and Q-Q plots. To address skewed distributions and enhance the interpretability of results, NT-proBNP, HALP and mHALP scores were log-transformed.

Univariate logistic regression models were performed to evaluate the association between HALP and mHALP with mortality. The results were presented as odds ratios (OR) with 95% confidence intervals (CI). The optimal cutoff values for HALP and mHALP were determined using Youden’s Index to maximize sensitivity and specificity. Univariate logistic regression was also conducted for other clinical variables, and those with a p-value <0.2 were considered as potential covariates in the multivariable analysis. Four stepwise multivariable logistic regression models, using backward selection with Akaike Information Criterion (AIC) used to balance model complexity and goodness-of-fit, were constructed to assess the independent effect of HALP, mHALP, and their binary versions based on Youden’s Index cutoff. Multicollinearity was assessed using the Variance Inflation Factor (VIF), and variables with a VIF > 10 were excluded or combined to prevent overfitting. All models’ performance was evaluated using Receiver Operating Characteristic (ROC) analysis, with the area under the curve (AUC) serving as the primary metric of discrimination. To address the class imbalance between survivors and non-survivors, we applied inverse probability weighting in all regression models, assigning greater weight to the minority class (90-day non-survivors) to ensure unbiased estimation of model coefficients. Model performance metrics (e.g., AUC) were optimism-adjusted using 1,000 bootstrap resamples.

All statistical analyses were conducted using R version 4.3. Model diagnostics, including residual analysis, influence statistics, and goodness-of-fit tests, were systematically examined to ensure model validity. All statistical tests were two-tailed, and a p-value < 0.05 was considered statistically significant.

## Results

A total of 1000 participants with CHF were enrolled in this study, of which 868 (86.8%) patients survived beyond 90 days, and 132 (13.2%) patients died within 90 days. The median age of the survivors was 71 (18.0) and that of the non-survivors was 77 (13.0). The surviving group of patients had a median HALP score of 27.18 (28.43) and the non-survivors had a median HALP score of 19.69 (21.39). The median mHALP scores were lower in non-survivors, 583,920.75 (940,245.74) compared to the survivors, 1,151,898.94 (1,293,723.58). Log-transformed HALP and mHALP scores naturally followed similar trends. Laboratory parameters including hemoglobin, albumin, absolute lymphocyte counts, and platelets were higher in the survivors’ group. NT-proBNP was significantly higher in the non-survivors, indicating poor cardiac function (7648.5 vs. 2393.0, p<0.001). There was a higher prevalence of advanced chronic kidney disease (CKD stage 4) in the non-survivor group (32.6%, p<0.001) than in the survivor group (16.9%). They were also more likely to suffer from acute renal failure (ARF), cardiac arrest, and shock (p<0.001).

In terms of medications, survivors were more likely to be prescribed ACE inhibitors (30.5% vs. 15.9%, p<0.001), ARBs (24.1% vs. 9.8%, p<0.001), beta-blockers (78.1% vs. 59.8%, p<0.001), and CCBs (19.9% vs. 8.3%, p=0.002). On the other hand, non-survivors were more frequently prescribed mineralocorticoid receptor antagonist (24.0% vs. 14.4%, p=0.020), amiodarone (28.0% vs. 16.6%, p=0.002), and vasopressors (52.3% vs. 15.9%, p<0.001). Non-survivors were also more likely to require invasive ventilation (33.3% vs. 10.5%, p<0.001) and dialysis (13.6% vs. 0.9%, p<0.001). Race and marital status were not significantly associated with mortality. Further, it was observed that patients with reduced ejection fraction (EF) were more susceptible to mortality compared to those with mildly reduced or preserved EF. Other significant baseline characteristics of the patients, stratified by 90 day mortality status, are presented in Table 1.

**Table 1.**
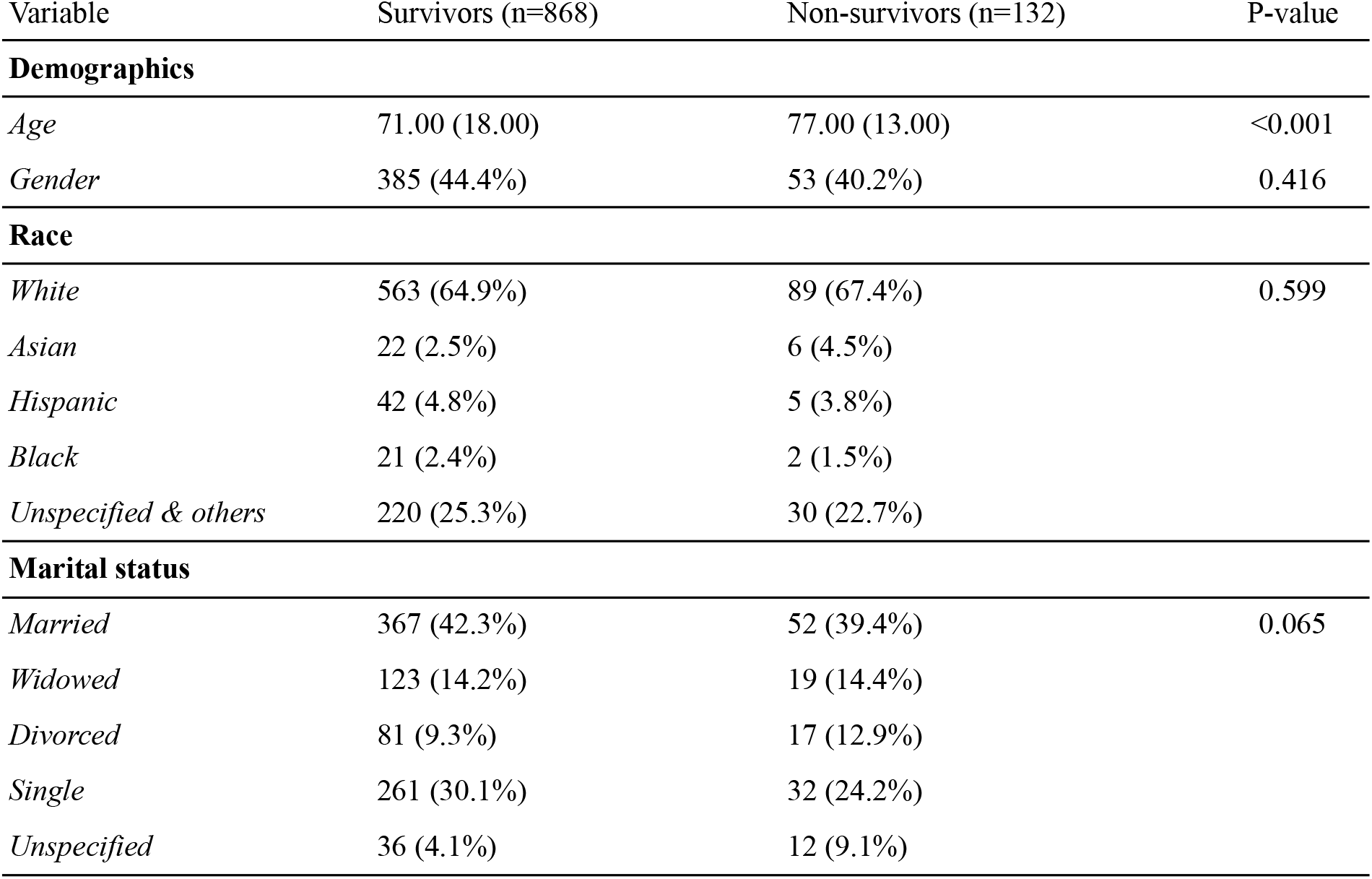
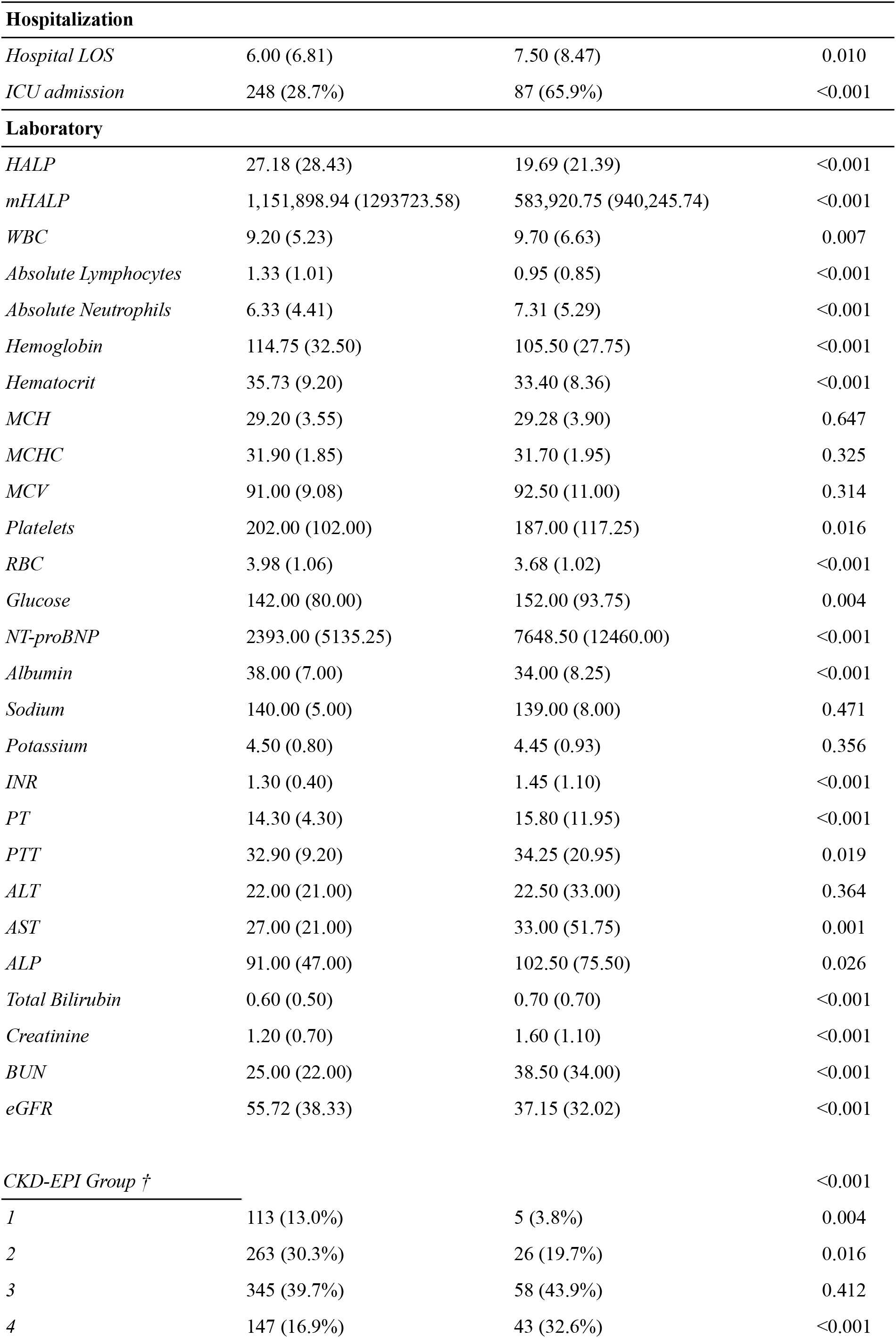
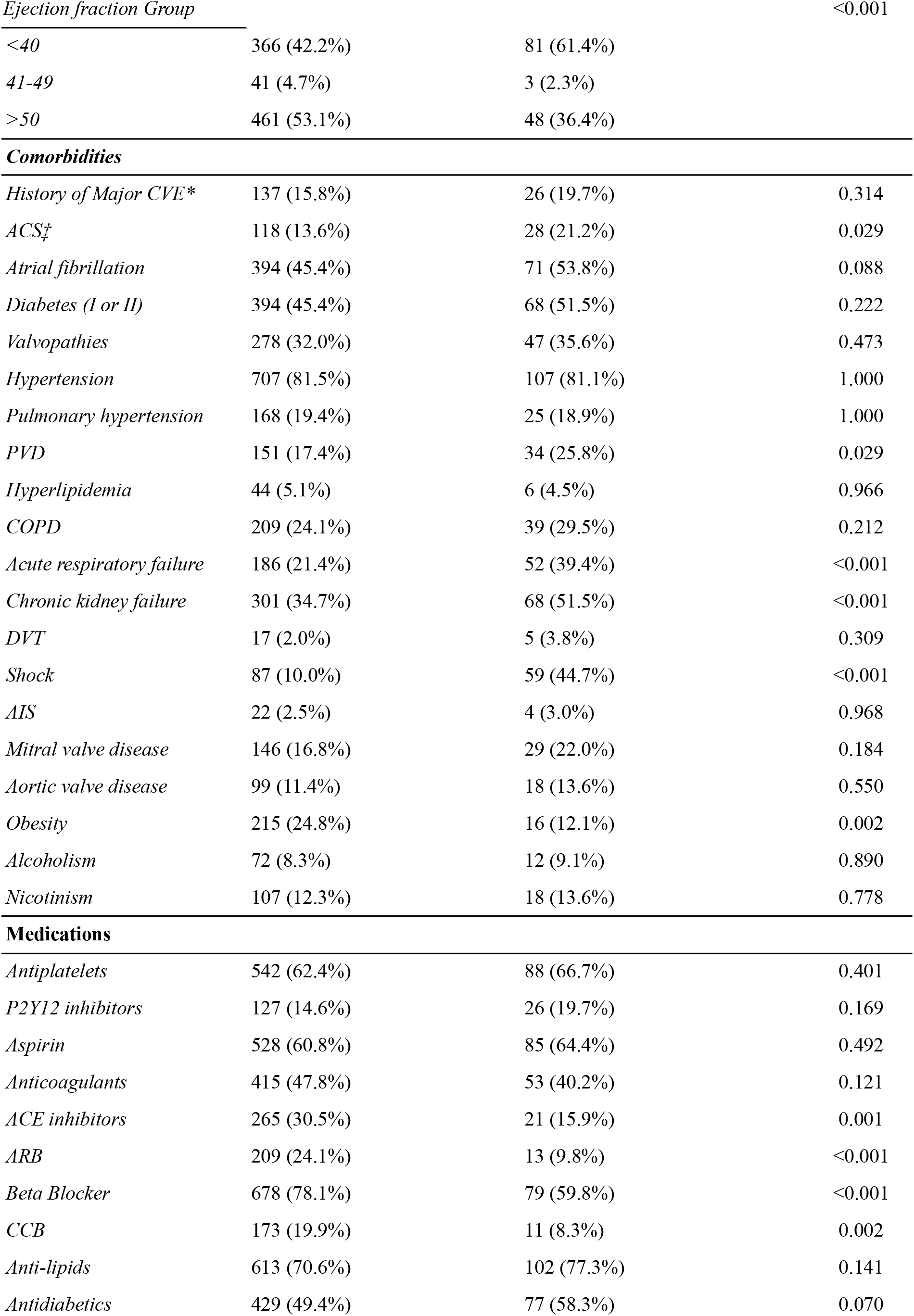
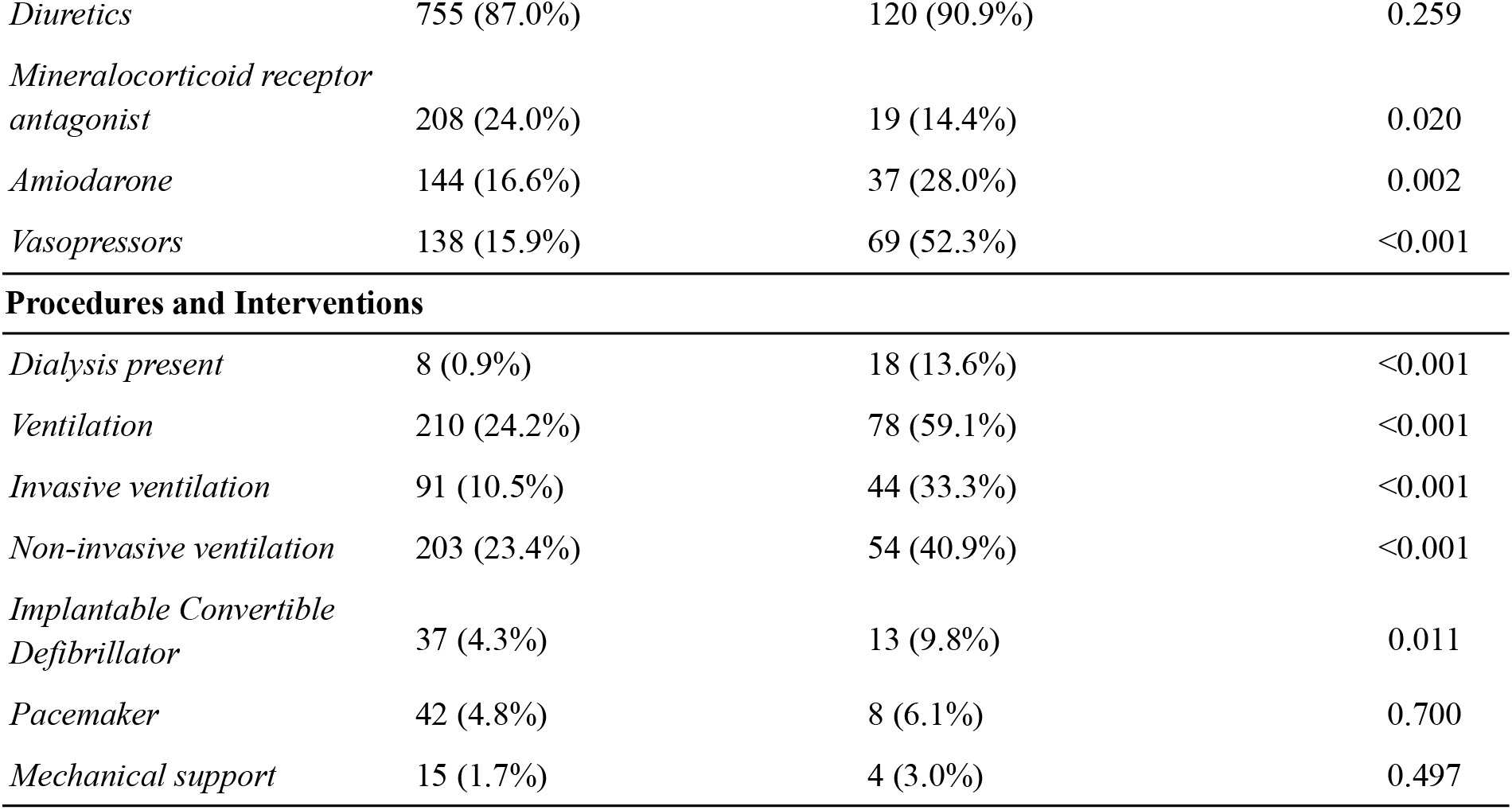
Baseline characteristics of patients stratified by 90-day mortality status. Continuous variables are presented as median (interquartile range), and categorical variables as number, percentage). P-values indicate the statistical significance of differences between survivors and non-survivors. †: Calculated using the 2021 formula. *:Previous MI, PE, AIS, or cardiac arrest. ‡: Defined as MI or unstable angina as per ESC guidelines on ACS management 2023.

| Variable | Survivors (n=868) | Non-survivors (n=132) | P-value |
| --- | --- | --- | --- |
| Demographics |  |  |  |
| Age | 71.00 (18.00) | 77.00 (13.00) | <0.001 |
| Gender | 385 (44.4%) | 53 (40.2%) | 0.416 |
| Race |  |  |  |
| White | 563 (64.9%) | 89 (67.4%) | 0.599 |
| Asian | 22 (2.5%) | 6 (4.5%) |  |
| Hispanic | 42 (4.8%) | 5 (3.8%) |  |
| Black | 21 (2.4%) | 2 (1.5%) |  |
| Unspecified & others | 220 (25.3%) | 30 (22.7%) |  |
| Marital status |  |  |  |
| Married | 367 (42.3%) | 52 (39.4%) | 0.065 |
| Widowed | 123 (14.2%) | 19 (14.4%) |  |
| Divorced | 81 (9.3%) | 17 (12.9%) |  |
| Single | 261 (30.1%) | 32 (24.2%) |  |
| Unspecified | 36 (4.1%) | 12 (9.1%) |  |

**Table 1.** Baseline characteristics of patients stratified by 90-day mortality status.
| **Hospitalization** |  |  |  |
| --- | --- | --- | --- |
| *Hospital LOS* | 6.00 (6.81) | 7.50 (8.47) | 0.010 |
| *ICU admission* | 248 (28.7%) | 87 (65.9%) | <0.001 |
| **Laboratory** |  |  |  |
| *HALP* | 27.18 (28.43) | 19.69 (21.39) | <0.001 |
| *mHALP* | 1,151,898.94 (1293723.58) | 583,920.75 (940,245.74) | <0.001 |
| *WBC* | 9.20 (5.23) | 9.70 (6.63) | 0.007 |
| *Absolute Lymphocytes* | 1.33 (1.01) | 0.95 (0.85) | <0.001 |
| *Absolute Neutrophils* | 6.33 (4.41) | 7.31 (5.29) | <0.001 |
| *Hemoglobin* | 114.75 (32.50) | 105.50 (27.75) | <0.001 |
| *Hematocrit* | 35.73 (9.20) | 33.40 (8.36) | <0.001 |
| *MCH* | 29.20 (3.55) | 29.28 (3.90) | 0.647 |
| *MCHC* | 31.90 (1.85) | 31.70 (1.95) | 0.325 |
| *MCV* | 91.00 (9.08) | 92.50 (11.00) | 0.314 |
| *Platelets* | 202.00 (102.00) | 187.00 (117.25) | 0.016 |
| *RBC* | 3.98 (1.06) | 3.68 (1.02) | <0.001 |
| *Glucose* | 142.00 (80.00) | 152.00 (93.75) | 0.004 |
| *NT-proBNP* | 2393.00 (5135.25) | 7648.50 (12460.00) | <0.001 |
| *Albumin* | 38.00 (7.00) | 34.00 (8.25) | <0.001 |
| *Sodium* | 140.00 (5.00) | 139.00 (8.00) | 0.471 |
| *Potassium* | 4.50 (0.80) | 4.45 (0.93) | 0.356 |
| *INR* | 1.30 (0.40) | 1.45 (1.10) | <0.001 |
| *PT* | 14.30 (4.30) | 15.80 (11.95) | <0.001 |
| *PTT* | 32.90 (9.20) | 34.25 (20.95) | 0.019 |
| *ALT* | 22.00 (21.00) | 22.50 (33.00) | 0.364 |
| *AST* | 27.00 (21.00) | 33.00 (51.75) | 0.001 |
| *ALP* | 91.00 (47.00) | 102.50 (75.50) | 0.026 |
| *Total Bilirubin* | 0.60 (0.50) | 0.70 (0.70) | <0.001 |
| *Creatinine* | 1.20 (0.70) | 1.60 (1.10) | <0.001 |
| *BUN* | 25.00 (22.00) | 38.50 (34.00) | <0.001 |
| *eGFR* | 55.72 (38.33) | 37.15 (32.02) | <0.001 |
| *CKD-EPI Group †* |  |  | <0.001 |
| *1* | 113 (13.0%) | 5 (3.8%) | 0.004 |
| *2* | 263 (30.3%) | 26 (19.7%) | 0.016 |
| *3* | 345 (39.7%) | 58 (43.9%) | 0.412 |
| *4* | 147 (16.9%) | 43 (32.6%) | <0.001 |
| *Ejection fraction Group* |  |  | <0.001 |
| <40 | 366 (42.2%) | 81 (61.4%) |  |
| 41-49 | 41 (4.7%) | 3 (2.3%) |  |
| >50 | 461 (53.1%) | 48 (36.4%) |  |
| ***Comorbidities*** |  |  |  |
| *History of Major CVE\** | 137 (15.8%) | 26 (19.7%) | 0.314 |
| *ACS‡* | 118 (13.6%) | 28 (21.2%) | 0.029 |
| *Atrial fibrillation* | 394 (45.4%) | 71 (53.8%) | 0.088 |
| *Diabetes (I or II)* | 394 (45.4%) | 68 (51.5%) | 0.222 |
| *Valvopathies* | 278 (32.0%) | 47 (35.6%) | 0.473 |
| *Hypertension* | 707 (81.5%) | 107 (81.1%) | 1.000 |
| *Pulmonary hypertension* | 168 (19.4%) | 25 (18.9%) | 1.000 |
| *PVD* | 151 (17.4%) | 34 (25.8%) | 0.029 |
| *Hyperlipidemia* | 44 (5.1%) | 6 (4.5%) | 0.966 |
| *COPD* | 209 (24.1%) | 39 (29.5%) | 0.212 |
| *Acute respiratory failure* | 186 (21.4%) | 52 (39.4%) | <0.001 |
| *Chronic kidney failure* | 301 (34.7%) | 68 (51.5%) | <0.001 |
| *DVT* | 17 (2.0%) | 5 (3.8%) | 0.309 |
| *Shock* | 87 (10.0%) | 59 (44.7%) | <0.001 |
| *AIS* | 22 (2.5%) | 4 (3.0%) | 0.968 |
| *Mitral valve disease* | 146 (16.8%) | 29 (22.0%) | 0.184 |
| *Aortic valve disease* | 99 (11.4%) | 18 (13.6%) | 0.550 |
| *Obesity* | 215 (24.8%) | 16 (12.1%) | 0.002 |
| *Alcoholism* | 72 (8.3%) | 12 (9.1%) | 0.890 |
| *Nicotinism* | 107 (12.3%) | 18 (13.6%) | 0.778 |
| ***Medications*** |  |  |  |
| *Antiplatelets* | 542 (62.4%) | 88 (66.7%) | 0.401 |
| *P2Y12 inhibitors* | 127 (14.6%) | 26 (19.7%) | 0.169 |
| *Aspirin* | 528 (60.8%) | 85 (64.4%) | 0.492 |
| *Anticoagulants* | 415 (47.8%) | 53 (40.2%) | 0.121 |
| *ACE inhibitors* | 265 (30.5%) | 21 (15.9%) | 0.001 |
| *ARB* | 209 (24.1%) | 13 (9.8%) | <0.001 |
| *Beta Blocker* | 678 (78.1%) | 79 (59.8%) | <0.001 |
| *CCB* | 173 (19.9%) | 11 (8.3%) | 0.002 |
| *Anti-lipids* | 613 (70.6%) | 102 (77.3%) | 0.141 |
| *Antidiabetics* | 429 (49.4%) | 77 (58.3%) | 0.070 |
| *Diuretics* | 755 (87.0%) | 120 (90.9%) | 0.259 |
| *Mineralocorticoid receptor antagonist* | 208 (24.0%) | 19 (14.4%) | 0.020 |
| *Amiodarone* | 144 (16.6%) | 37 (28.0%) | 0.002 |
| *Vasopressors* | 138 (15.9%) | 69 (52.3%) | <0.001 |
| **Procedures and Interventions** |  |  |  |
| *Dialysis present* | 8 (0.9%) | 18 (13.6%) | <0.001 |
| *Ventilation* | 210 (24.2%) | 78 (59.1%) | <0.001 |
| *Invasive ventilation* | 91 (10.5%) | 44 (33.3%) | <0.001 |
| *Non-invasive ventilation* | 203 (23.4%) | 54 (40.9%) | <0.001 |
| *Implantable Convertible Defibrillator* | 37 (4.3%) | 13 (9.8%) | 0.011 |
| *Pacemaker* | 42 (4.8%) | 8 (6.1%) | 0.700 |
| *Mechanical support* | 15 (1.7%) | 4 (3.0%) | 0.497 |

Univariate logistic regression analysis of log-transformed HALP and mHALP scores for 90-day mortality in patients with CHF is presented in Table 2. The AUC-ROC curves are provided in Figure 1. The regression analysis for HALP revealed an area under the curve (AUC) of 0.63 (95% CI: 0.60–0.66) and an odds ratio (OR) of 0.25 (95% CI: 0.14 - 0.44). The optimal cut-off value for HALP was determined to be 1.20 with a sensitivity of 73.0% and specificity of 46.2%. On the other hand, mHALP had a higher AUC 0.68 (95% CI: 0.65–0.72) and the strongest negative association with 90 day mortality, with an OR of 0.22 (95% CI: 0.14–0.35). The optimal cut-off value for mHALP was 5.82, with sensitivity and specificity at 72.1% and 56.1%, respectively.

**Table 2.** Crude logistic regression analyses for log-transformed HALP and mHALP. Optimal cutoff obtained using Youden’s Index.

| Biomarker | OR (95%CI) | P-value | AUC (95%CI) | Optimal cutoff | Sensitivity | Specificity |
| --- | --- | --- | --- | --- | --- | --- |
| *HALP* | 0.25 (0.14, 0.44) | <0.001 | 0.63 (0.60-0.66) | 1.20 | 73.0% | 46.0% |
| *mHALP* | 0.22 (0.14, 0.35) | <0.001 | 0.68 (0.65-0.72) | 5.82 | 72.1% | 56.0% |

**Figure 1.**
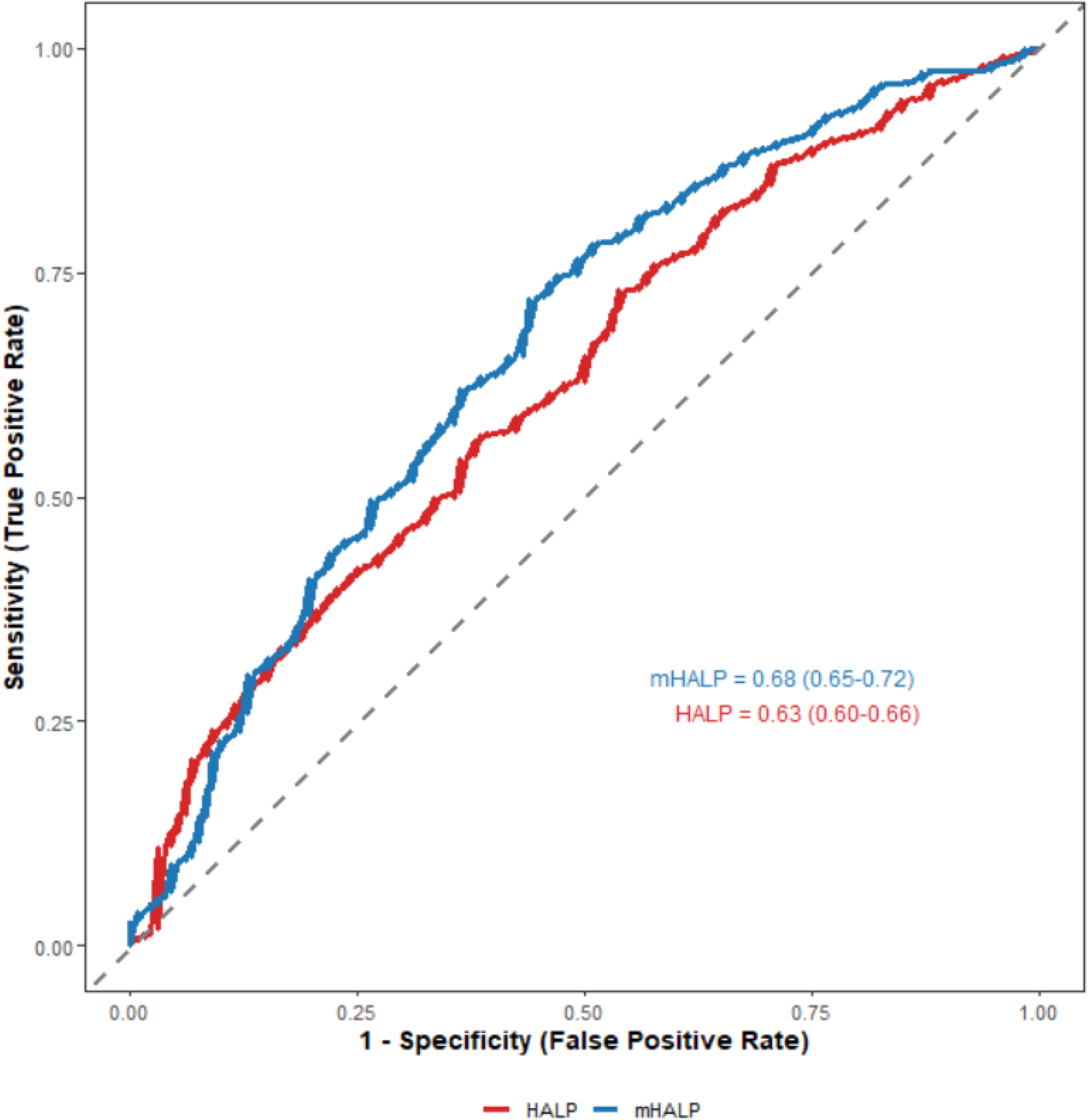
ROC curves for HALP and mHALP in predicting 90-day mortality.

Table 3 shows the multivariate logistic regression analyses for HALP and mHALP, adjusted for potential confounders. Four distinct models—HALP, mHALP, and their binary representations —were independently analyzed to assess the association of each variable with 90 day mortality. The models were adjusted for age, EF categories, ICU stay, ACEI/ARBs, beta-blockers, CCBs, antidiabetic medications, vasopressors, shock, implantable cardioverter-defibrillator and NT-proBNP levels. Both HALP (OR: 0.44; 95% CI: 0.22–0.87) and mHALP (OR: 0.57; 95% CI: 0.32–0.86) were found to be significant predictors of 90 day mortality.

**Table 3.**
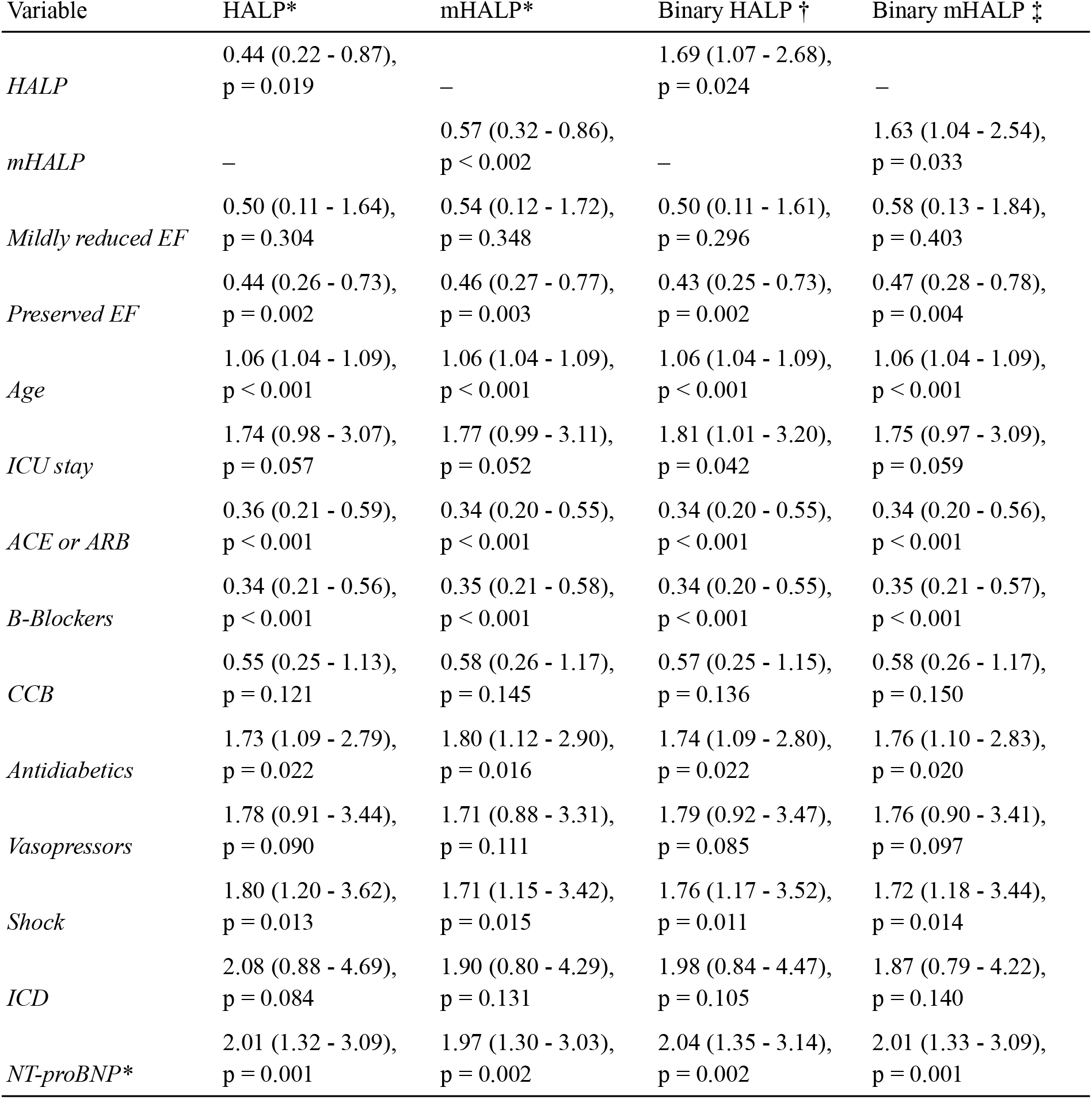
Final stepwise backward regression model, with 90-dat mortality as outcome. *:Log-transformed. †: High HALP (≥1.20) as reference. ‡: High mHALP (≥5.82) as reference. ICD: Implantable cardioverter-defibrillator.

Among the covariates, age was a strong predictor of mortality in all models. Covariates such as ICU stay and NT-proBNP levels were identified as major independent risk factors for 90 day mortality in CHF patients. While the use of antidiabetic drugs (p<0.05) was associated with an increased risk of mortality, ACEI/ARBs and beta-blockers were significantly associated with reduced mortality risk.

Kaplan-Meier survival analysis was done to compare the probability of 90 day survival between the two groups, stratified by HALP and mHALP scores (Figure 2). Both curves demonstrate that patients with high HALP and mHALP scores have a higher survival probability than patients with low scores (p<0.001).

**Figure 2.**
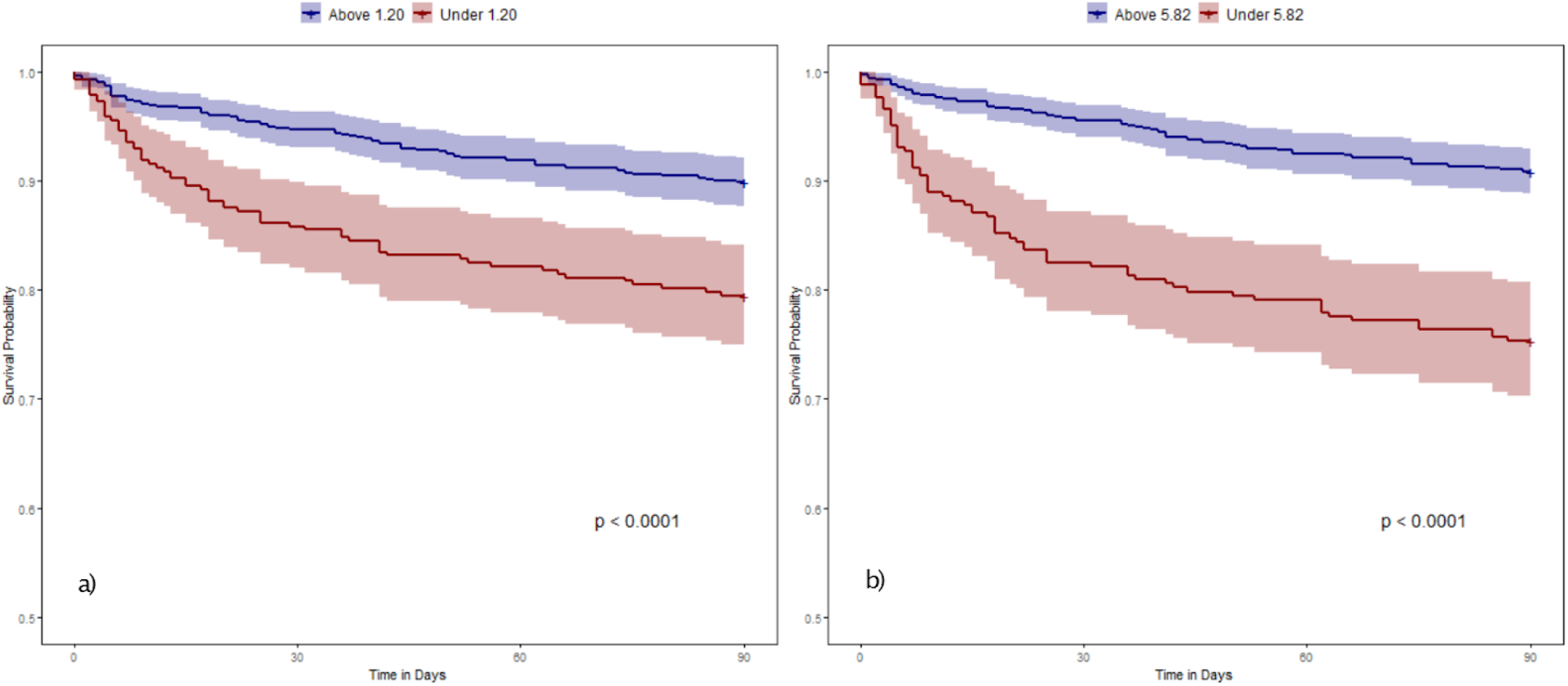
90-day survival Kaplan-Meier curves for a) HALP and b) mHALP scores stratified by optimal cutoffs.

## Discussion

This study evaluated the predictive value of HALP and mHALP on 90-day mortality in 1000 CHF patients admitted to the hospital. The non-survivor group had lower median HALP and mHALP scores compared to the survivors (p<0.001) in both crude and adjusted regression analyses. Both scores had good discriminatory values; nonetheless, mHALP showed better performance across all metrics, standing out as a promising predictor of mortality in CHF patients.

The HALP score has gained significant traction in oncology, as it effectively reflects the immuno-nutritional status of cancer patients and shows the potential to predict prognosis and mortality [10]. Inflammation and nutrition play a key role in the pathophysiology of CHF and are independently associated with mortality [17,18]. Hemoglobin synthesis depends on adequate levels of nutrients obtained from the diet (e.g., iron, vitamin B12). Low hemoglobin levels indicate anemia, which exacerbates tissue hypoxia and increases cardiac workload. Albumin, a negative acute-phase reactant protein, is decreased in states of inflammation and malnutrition. Hypoalbuminemia reduces oncotic pressure, leading to fluid leakage and exacerbating CHF symptoms such as edema. Lymphocytes are a fundamental part of the body’s immune response, and low counts reflect immune dysfunction and chronic inflammation, which contribute to CHF progression. Platelets play a key role in hemostasis and tissue repair. Abnormal platelet counts (thrombocytosis and thrombocytopenia) increase the risk of thromboembolism and bleeding complications, which are linked with higher mortality. Together, these biomarkers make the HALP score a valuable tool for assessing patients’ hematological, inflammatory, and nutritional status and predicting mortality in CHF patients. Chronic inflammation exacerbates CHF progression by promoting oxidative stress, a state of imbalance between reactive oxygen species (ROS) production and antioxidant defenses [26]. This oxidative stress damages cardiomyocytes and contributes to myocardial dysfunction [27]. Furthermore, both inflammation and malnutrition can disrupt neurohormonal regulation, leading to overactivation of the RAAS and sympathetic nervous system. In turn, this causes increased vasoconstriction, sodium and water retention, and further cardiac workload [28]. Along with direct effects of inflammatory mediators on the myocardium, these factors accelerate adverse cardiac remodeling, characterized by fibrosis, hypertrophy, and ultimately, reduced contractile function [29].

In the original article, HALP score was confirmed as an independent prognostic factor of gastric carcinoma, and patients with low HALP scores had lower 1, 2, and 3-year survival rates compared to the high HALP group [11]. Similar results were observed in studies conducted in other clinical settings, suggesting that low HALP scores indicate short- and long-term mortality [19-23]. These findings validate the results of this study, which found that survivors had higher median HALP and mHALP scores than the non-survivors. A retrospective study consisting of 730 patients compared the predictive ability of HALP with the MAGGIC score, C-reactive protein (CRP), and prognostic nutritional index (PNI) [24], finding that HALP had higher AUC than the other indexes for predicting 1-month (0.70, 95% CI: 0.63–0.76)) and 1-year (0.68, 95% CI: 0.62–0.74) mortality, indicating that the HALP score is a good predictor of mortality in CHF patients. Another study compared the diagnostic performance of the individual parameters of HALP with the HALP score in predicting short-term mortality among acute decompensated CHF patients and found that albumin had the highest AUC (0.81) followed by HALP score (AUC: 0.77) [25]. The high AUC value demonstrates that HALP can be considered a reliable prognostic tool for mortality in acute CHF settings. However, the performance observed in our analysis was significantly lower (AUC = 0.63). We attribute this difference to methodological variations, including the earlier study’s endpoint (in-hospital mortality), smaller sample (n=227), and limited variable testing potentially inflating performance estimates. A study conducted on 101 acute CHF patients and found that the classical HALP score was not a significant predictor of 1-week and 3-month mortality [15]. According to the original HALP score’s formula, hemoglobin, albumin, and lymphocytes are divided by platelet count, which may disproportionately lower the score predictive value in patients with thrombocytopenia, potentially masking the influence of the other components. To address and test this limitation, the authors developed a modified version of the HALP score, termed mHALP, where platelets are multiplied instead of divided, thus reflecting the impact of platelet count as a marker of inflammation and overall health status in patients. They found that mHALP was significantly associated with 3-month mortality.

Crude analysis revealed that HALP had an AUC of 0.63 (95% CI: 0.60–h0.66), indicating moderate predictive ability for 90 day mortality. Its strong association with mortality was reflected by an OR of 0.25 (0.14–0.44), with an optimal threshold of 1.20, which yielded a sensitivity of 73.0% and specificity of 46.2%. On the other hand, mHALP had better discriminatory power predicting 90-day mortality, with an AUC of 0.68 (95% CI: 0.65–0.72) and an OR of 0.22 (95% CI: 0.14–0.35). The optimal cut-off value of mHALP was 5.82, providing a sensitivity of 72.1% and specificity of 56.1%. Compared to HALP, mHALP demonstrated superior predictive ability and higher specificity, emphasizing its discriminatory power in distinguishing between high-risk patients and those with a better prognosis. The relatively high sensitivity and lower specificity suggest that these scores may be an effective screening tool for identifying patients at high risk of mortality. The moderate AUC values of HALP and mHALP suggest that while these scores are effective, combining them with other clinical risk models would improve the overall predictive power of mortality in CHF patients.

In multivariate analyses, both the continuous and binary forms of HALP and mHALP scores were significantly associated with reduced mortality, suggesting their utility as effective tools for risk stratification. Age, shock, and log-transformed NT-proBNP emerged as independent predictors of 90 day mortality in CHF, highlighting their value in predictive models and risk scores. The use of antidiabetic medications, particularly insulin, being the most commonly prescribed, was associated with an increased mortality risk. This association likely reflects the underlying severity of diabetes or the presence of diabetes-related complications, rather than a direct effect of the medications itself. Diabetes is strongly linked to inflammation and endothelial damage, driven by hyperglycemia, oxidative stress and metabolic disturbances [30]. Supporting this, a large-scale study of 21,578 participants, which identified the HALP score as an independent predictor of mortality, found diabetes to be associated with all-cause mortality (p=0.03) [22]. In the context of our findings, the increased mortality risk associated with antidiabetic use should be interpreted as a marker of disease severity instead of a direct risk of mortality. The protective effects of ACEi, ARBs and beta-blockers observed in this analysis are consistent with current treatment guidelines for CHF and their role in reducing mortality [1]. In the analysis, preserved EF was significantly associated with reduced mortality across all models compared to the reference category of reduced EF. In contrast, mildly reduced EF showed a non-significant association with mortality. The lower mortality observed in patients with preserved EF aligns with the generally better prognosis associated with this subgroup, likely due to less severe systolic dysfunction and a distinct underlying pathophysiology compared to those with reduced EF. Implantable cardioverter-defibrillator and cardiogenic shock showed trends toward increased mortality risk, reflecting the severity of CHF and cardiac dysfunction in these patients. Overall, these findings point to the significant predictive ability of HALP and mHALP scores after adjusting for other covariates, while also underscoring the value of further investigating variables such as age and NT-proBNP to potentially enhance the HALP and mHALP equations for improved mortality risk assessment in this cohort.

Both the crude and adjusted analyses demonstrated the strong predictive performance of HALP and mHALP scores and their significant association with mortality. The results of this study align with previous research, reinforcing the reliability of these scores, especially mHALP, as a robust predictor of 90-day mortality in CHF patients admitted to the hospital.

### Clinical Utility

The HALP and mHALP scores offer a cost-effective, accessible, and straightforward approach to predicting mortality in CHF patients. Derived from routine laboratory parameters, these scores can be calculated upon hospital admission at minimal expense, providing clinicians with an immediate snapshot of a patient’s mortality risk. Their simplicity and reliance on widely available tests make them particularly valuable in resource-limited settings, where advanced diagnostic tools may be scarce. Moreover, the dynamic nature of these scores enables serial measurements throughout a patient’s hospital stay, allowing physicians to monitor disease progression and adjust treatment strategies as needed. However, it is important to note that the moderate AUC and low specificity observed in this study suggest that these scores are best used to supplement, rather than replace, existing models or clinical judgement. Therefore, integrating HALP and mHALP with established predictive tools—such as the Seattle Heart Failure Model or MAGGIC Risk Score—could further enhance risk stratification, improving the precision of mortality predictions and supporting tailored clinical decision-making in CHF management.

### Limitations

Several limitations should be considered when interpreting the findings of this study. First, the retrospective design, while allowing for analysis of a large dataset, is susceptible to inherent biases, including potential selection bias and reliance on routinely collected electronic health record (EHR) data accuracy. Causal inferences cannot be definitively drawn from this observational data. Second, although the MIMIC-IV database provides a substantial sample size (n=1000), the data originate from a single, large academic medical center (Beth Israel Deaconess Medical Center). Therefore, caution should be exercised when generalizing these findings to potentially different patient populations encountered in other settings, such as community hospitals or diverse geographical regions. Third, our methodology required complete clinical and laboratory data for patient inclusion. While this ensures uniformity for the calculation of HALP/mHALP and adjustment variables, this complete case analysis approach could potentially introduce selection bias if the characteristics of patients with missing data differ systematically from those included. Fourth, while we employed multivariate logistic regression to adjust for a wide range of potential confounders, the possibility of residual confounding from unmeasured variables (such as socioeconomic factors, detailed functional status, medication adherence, or specific etiologies of CHF not captured in routine data) cannot be entirely excluded. Finally, the HALP and mHALP scores demonstrated moderate discriminative ability for 90-day mortality in this cohort (AUC 0.63 and 0.68, respectively), with relatively modest specificity. This suggests that while statistically significant and potentially useful, these scores should be considered as supplementary tools to aid risk stratification alongside comprehensive clinical evaluation and established prognostic models, rather than as standalone replacements. Future prospective, multi-center studies are warranted to validate these findings and further explore the integration of these scores into clinical practice across broader populations.

## Conclusion

This study demonstrates that both the HALP and mHALP scores are significantly associated with 90-day mortality in patients with congestive heart failure (CHF). Notably, the modified mHALP score exhibited a superior predictive performance compared to the traditional HALP score. These findings suggest that routine calculation of these scores using readily available laboratory parameters may offer a cost-effective and rapid method for risk stratification in CHF. By identifying high-risk patients early in their hospital admission, clinicians may be better equipped to tailor interventions and optimize management strategies. Future research should focus on further refining these indices—potentially incorporating additional prognostic variables such as age, ejection fraction, and biomarkers like NT-proBNP—to enhance predictive accuracy and validate their utility across larger, multicenter populations.

## Data Availability

All data produced in the present study are available upon reasonable request to the authors.

https://physionet.org/content/mimiciv/

## Abbreviation

ACE inhibitor: Angiotensin-converting enzyme inhibitor
ACS: Acute coronary syndrome
AIS: Acute ischemic stroke
ALT: Alanine transaminase
ALP: Alkaline phosphatase
ARB: Angiotensin receptor blocker
ARF: Acute renal failure
BUN: Blood urea nitrogen
CCB: Calcium channel blocker
CHF: Congestive heart failure
CKD-EPI: Chronic Kidney Disease Epidemiology Collaboration
COPD: Chronic obstructive pulmonary disease
CRP: C-reactive protein
EF: Ejection Fraction
eGFR: Estimated glomerular filtration rate
HALP score: Hemoglobin albumin lymphocyte platelet score
INR: International normalized ratio
LOS: Length of stay
Major CVE: Major cardiovascular event
MCH: Mean corpuscular hemoglobin
MCHC: Mean corpuscular hemoglobin concentration
MCV: Mean corpuscular volume
mHALP score: Modified hemoglobin albumin lymphocyte platelet score
NT-proBNP: N-terminal pro b-type natriuretic peptide
PNI: Prognostic nutritional index
PT: Prothrombin time
PTT: Partial thromboplastin time
PVD: Peripheral vascular disease
RBC: Red blood cells
WBC: White blood cells

